# Consensus Guideline for the Management of Malignant Gastrointestinal Obstruction in Patients with Peritoneal Surface Malignancies

**DOI:** 10.1101/2024.04.09.24305427

**Authors:** PSM Writing Group, PSM Consortium Group, Kiran K. Turaga

## Abstract

**Background:** Malignant gastrointestinal obstruction (MGIO), a frequent complication of peritoneal surface malignancies (PSM), often portends a poor prognosis. The lack of high-quality evidence on optimal management strategies necessitated a national consensus to address this clinical problem.

**Methods:** A clinical management pathway was designed through a Delphi consensus process with national experts in peritoneal disease. Two rounds of voting were conducted to assess agreement levels with pathway blocks. Supporting evidence regarding procedural interventions for MGIO underwent evaluation via a rapid literature review.

**Results:** Of 111 participants responding in the first round, 90 (81%) responded in the second round. Over 90% consensus was achieved in 4/6 and 6/6 pathway blocks during rounds I and II, respectively. Encouraging a multidisciplinary approach, the pathway emphasized early palliative care assessments and iterative goals of care evaluation throughout treatment. Management was delineated based on obstruction acuity, and selection criteria for palliative-intent surgical interventions and stenting were elucidated. Studies demonstrated limited benefits for such interventions in patients with multifocal obstructions, poor performance status, and high-grade and/or high-burden PSMs. In these cases, a recommendation for supportive care or upper GI decompression tube placement was favored. The overall level of evidence was generally low-moderate in existing literature.

**Conclusion:** Given limited evidence, the consensus-driven pathway provides crucial clinical guidance for practitioners dealing with MGIO in PSM patients. There is a need for high-quality prospective evidence in this domain.

## INTRODUCTION

Malignant gastrointestinal obstruction (MGIO) poses a significant challenge in the context of active intraperitoneal malignancy. Proposed mechanisms include extrinsic compression, adhesions, and cicatrization from peritoneal metastases, along with endoluminal obstructions in cases of primary gastrointestinal malignancies.^1^ Potential sites of obstruction include the large bowel and small bowel below the ligament of Treitz, referred to as malignant bowel obstruction (MBO), or proximal locations including the stomach and duodenum.^1^ This complication frequently occurs in patients with peritoneal surface malignancies (PSM), potentially affecting over 50% of individuals with gastrointestinal and genitourinary malignancies within 2-3 years of cancer diagnosis.^2–9^ MGIO is often indicative of advanced, unresectable disease and contributes substantially to mortality in patients with PSM owing to malnutrition, systemic therapy interruption, and acute sequelae like bowel perforation and bleeding.^10^ Hence, patients with MGIO-PSM comprise a vulnerable population with limited therapeutic options.

Two decades ago, an international conference highlighted challenges in palliative research for cancer patients. Participants cited ethical dilemmas for clinical trial enrollment, communication barriers, and unclear best practice standards as barriers to optimal trial design.^11^ Due to its prevalence, MGIO emerged as a promising model for researchers to study divergent treatment approaches and their impact on patient morbidity. Despite increased interest and advancements in patient management strategies, the literature guiding treatment for MGIO has remained relatively sparse and lends itself to variations based on individual and institutional experiences.^11,12^ For patients with MGIO, peritoneal carcinomatosis is consistently identified as a poor prognostic factor as these patients often present with multifocal, high grade, and angulated obstructions which limit the success of surgical and non-surgical interventions.^5,13–24^ A scarcity of high-quality comparative evidence has hindered the establishment of standard care management pathways for patients with MIGO caused by PSM.

Given the limited quality and heterogeneity of existing research, we aimed to create expert-based consensus recommendations regarding the approach to these complex patients. The goal is to formulate a clinical management pathway and synthesize evidence supported by the literature on surgical and procedural interventions for MGIO.

## METHODS

This initiative was part of a national multidisciplinary consortium group process aimed at streamlining guidelines for the care of patients with PSM. The consensus and rapid review methodology has been described in detail in a separate manuscript (Submitted).^25^ Major components are summarized below.

### Consensus Group Structure

In brief, the MGIO Working Group consisted of 11 representatives with expertise in surgical oncology, medical oncology, specialist palliative care, and nutrition. Two core group members (VVB and EG) coordinated the effort. A team of eight surgical residents, surgical oncology fellows, and research fellows conducted the rapid reviews.

### Modified Delphi Process

The 2018 Chicago Consensus Guidelines for PSM served as a foundation, offering consensus-based pathways for managing primary and secondary PSMs, along with a descriptive overview of palliative care considerations.^26^ However, a formal pathway for the management of MGIO was lacking.^27^ In this iteration, a pathway for MGIO management emerged through a modified Delphi consensus with two rounds of voting. Experts rated their agreement levels on a five-point Likert scale via a Qualtrics questionnaire. A 75% consensus threshold was set and blocks below 90% agreement underwent further review.

### Rapid Review of the Literature

A MEDLINE search via PubMed between January 2000 and August 2023 addressed the key question: ‘In patients with MGIO due to peritoneal carcinomatosis, are procedural interventions more or less effective and safe than medical therapy or observation alone?’ A search strategy was developed by co-author VVB and reviewed by a medical librarian specialist, and the review protocol was pre-registered in PROSPERO (<u>PROSPERO 2023 CRD42023463245</u>). The Covidence platform was used to facilitate title and abstract screening, full-text review, and data extraction. The extraction variables encompassed a thorough definition of study parameters, including patient characteristics, intervention attributes, and comparison groups if applicable. Relevant outcome measures included technical and clinical success rates, quality of life, initiation of post-intervention systemic therapy, recurrence of obstructive symptoms, post-intervention morbidity and mortality rates, and overall survival. Given that all included studies were non-randomized studies, quality assessment was performed using the Newcastle Ottawa Scale for cohort studies As described previously, a maximum of nine stars can be allotted across three quality domains: case selection, comparability of cohorts, and outcomes.^28,29^ Studies with six or higher stars are considered to be of high methodologic quality.^30^ The review was conducted in alignment with recommendations from the Cochrane Rapid Review Methods Groups and reported in line with the PRISMA 2020 guidelines.^31,32^

### External Perspectives

The guideline also incorporates international perspectives to evaluate global practice variability in the management of MGIO. For this purpose, the second version of the pathway was circulated amongst members of the Peritoneal Surface Oncology Group International (PSOGI) executive council. Their comments were consolidated along with a review of existing national and international guidelines regarding the management of MGIO to evaluate points of alignment and any major differences compared to our recommendations.

## RESULTS

### Pathways

In all, 111 experts voted on the clinical pathway during the first Delphi round. The responders included 85 (77%) surgical oncologists, 14 (13%) medical oncologists, 4 (3%) palliative care specialists, 3 (3%) pathologists, and 5 (4%) experts in other domains. Of these, 90 (81%) participants responded in the second round. Given the generally low-moderate quality of evidence in existing literature, most recommendations incorporated expert opinions. The pathway was divided into six main blocks (Figure 1), which are elucidated further.

**Figure 1:**
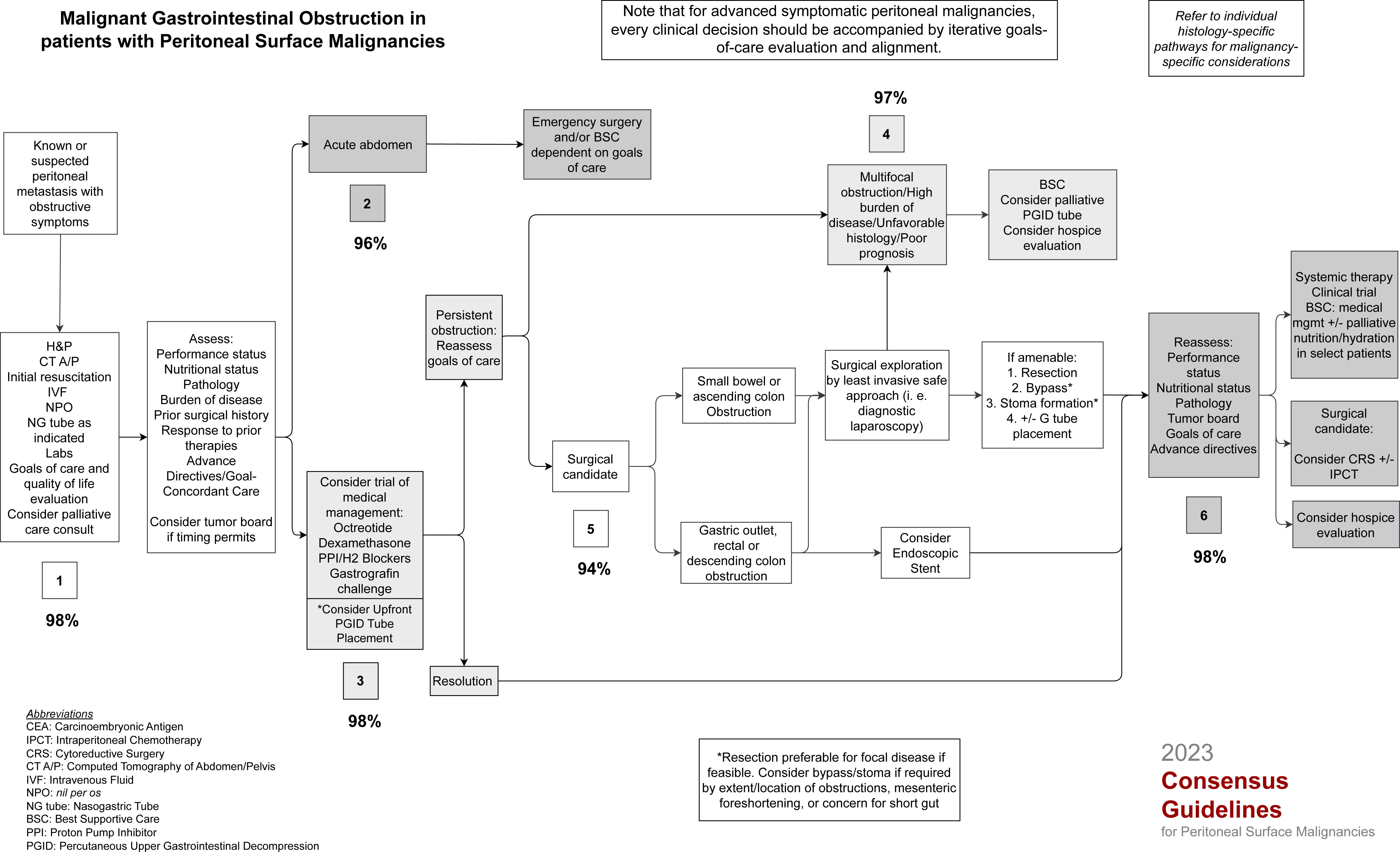
Pathway for the management of malignant gastrointestinal obstruction in patients with peritoneal surface malignancies.

### Rapid review

Of 1822 abstracts screened, 243 were included for full text review, and 25 studies reporting outcomes specific to patients with peritoneal metastases were included for data extraction and quality assessment. Of these, 12 studies focused on surgical palliation,^33–44^ seven studies focused on percutaneous upper GI decompression (PGID),^7–9,45–48^ and three studies each focused on upper GI and colorectal stenting.^17,18,23,49–51^ These studies are summarized in Tables 3-5 and elaborated upon in recommendations for blocks 2, 4 and 5. Relevant exclusion criteria are detailed in the PRISMA flow diagram (Figure 2). Of note, 39 studies were excluded as they did not report outcomes for the subgroup of patients with peritoneal metastases. While not summarized separately, some of these studies are referenced in relevant sections below.

**Figure 2:**
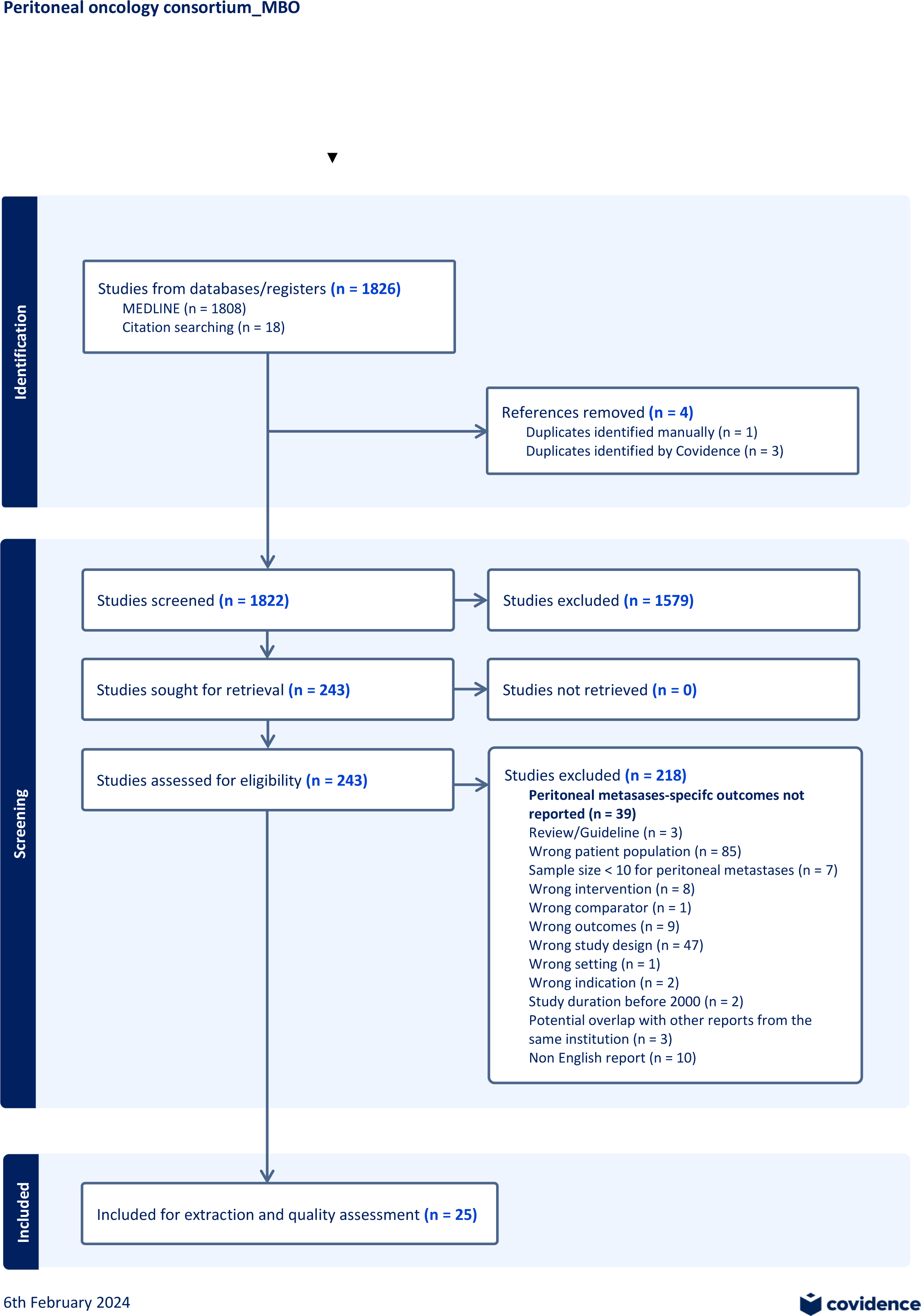
PRISMA flow diagram for the rapid review regarding surgical and procedural interventions for Malignant Gastrointestinal Obstruction in patients with Peritoneal Surface Malignancies.

**Tables 1 & 2:** Agreement tables for the two rounds of the modified delphi consensus. Percentage agreement corresponds to the proportion of responses marked as ‘Strongly Agree’ or ‘Agree’ per block.

Round I
|  | Strongly agree | Agree | Neither agree nor disagree | Disagree | Strongly disagree | Total | % Agree |
| --- | --- | --- | --- | --- | --- | --- | --- |
| **Block 1** | 83 | 25 | 2 | 1 | 0 | 111 | 97% |
| **Block 2** | 61 | 29 | 8 | 11 | 2 | 111 | 81% |
| **Block 3** | 69 | 35 | 5 | 2 | 0 | 111 | 94% |
| **Block 4** | 77 | 31 | 3 | 0 | 0 | 111 | 97% |
| **Block 5** | 62 | 33 | 10 | 6 | 0 | 111 | 86% |
| **Block 6** | 63 | 40 | 8 | 0 | 0 | 111 | 93% |

Round II
|  | Strongly agree | Agree | Neither agree nor disagree | Disagree | Strongly disagree | Total | % Agree |
| --- | --- | --- | --- | --- | --- | --- | --- |
| **Block 1** | 82 | 6 | 0 | 2 | 0 | 90 | 98% |
| **Block 2** | 80 | 6 | 2 | 2 | 0 | 90 | 96% |
| **Block 3** | 81 | 7 | 1 | 1 | 0 | 90 | 98% |
| **Block 4** | 80 | 7 | 2 | 1 | 0 | 90 | 97% |
| **Block 5** | 77 | 8 | 3 | 1 | 1 | 90 | 94% |
| **Block 6** | 81 | 7 | 1 | 1 | 0 | 90 | 98% |

**Table 3:**
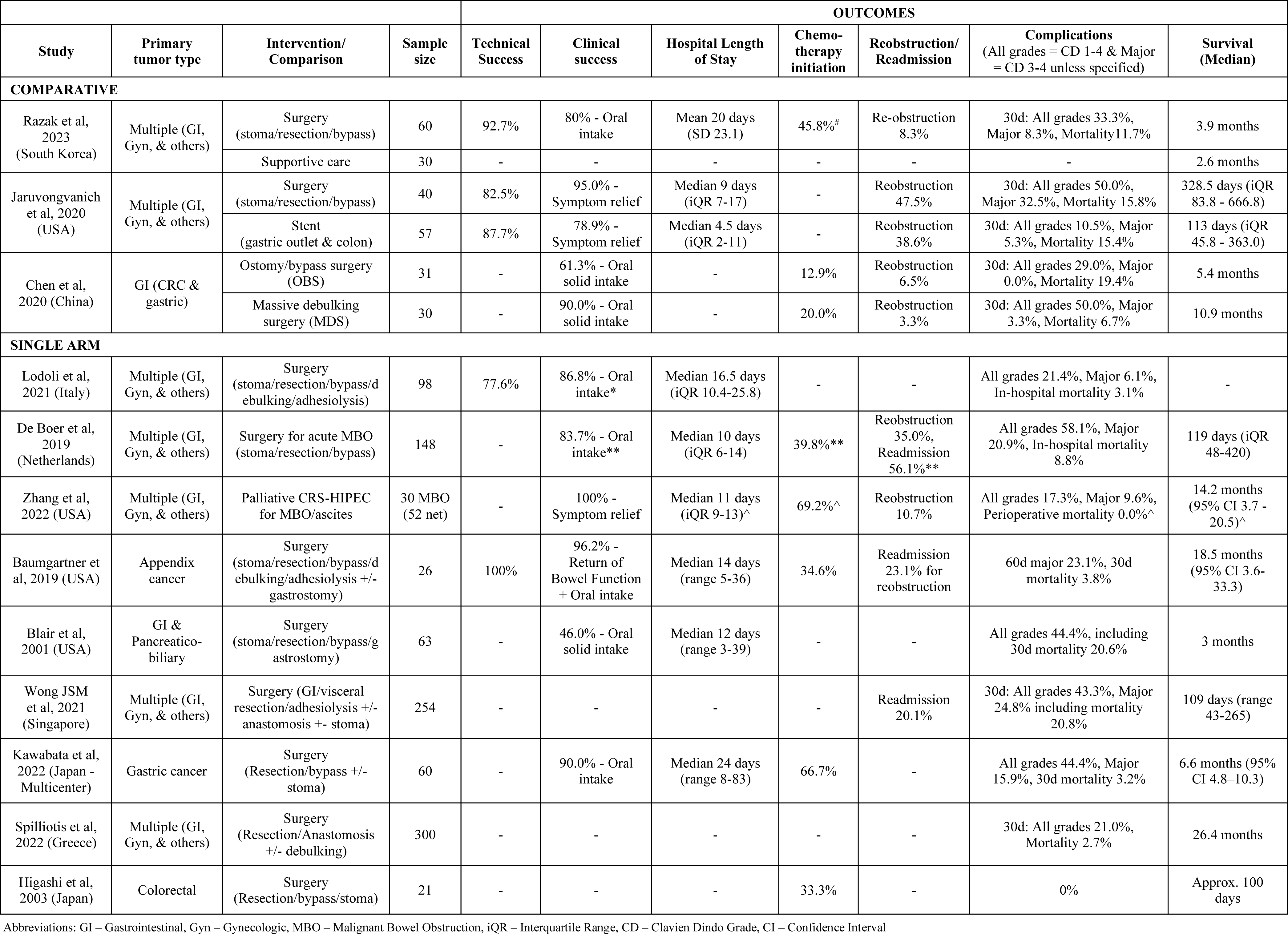
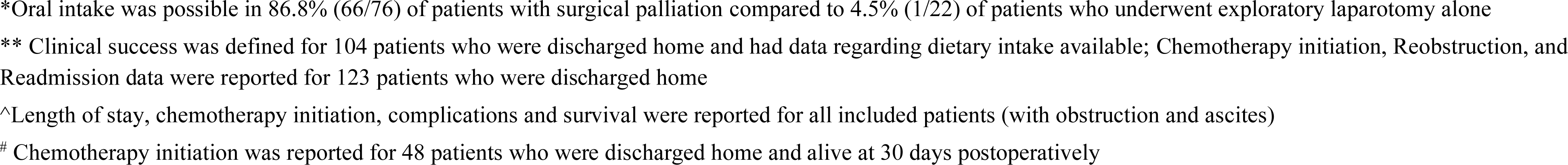
Summary of included studies focusing on surgical palliation of gastrointestinal obstruction in patients with peritoneal metastases.

|  |  |  |  | OUTCOMES |  |  |  |  |  |  |
| --- | --- | --- | --- | --- | --- | --- | --- | --- | --- | --- |
| Study | Primary tumor type | Intervention/ Comparison | Sample size | Technical Success | Clinical success | Hospital Length of Stay | Chemo-therapy initiation | Reobstruction/ Readmission | Complications (All grades = CD 1-4 & Major = CD 3-4 unless specified) | Survival (Median) |
| COMPARATIVE |  |  |  |  |  |  |  |  |  |  |
| Razak et al, 2023 (South Korea) | Multiple (GI, Gyn, & others) | Surgery (stoma/resection/bypass) | 60 | 92.7% | 80% - Oral intake | Mean 20 days (SD 23.1) | 45.8% # | Re-obstruction 8.3% | 30d: All grades 33.3%, Major 8.3%, Mortality 11.7% | 3.9 months |
|  |  | Supportive care | 30 | - | - | - | - | - | - | 2.6 months |
| Jaruvongvanich et al, 2020 (USA) | Multiple (GI, Gyn, & others) | Surgery (stoma/resection/bypass) | 40 | 82.5% | 95.0% - Symptom relief | Median 9 days (iQR 7-17) | - | Reobstruction 47.5% | 30d: All grades 50.0%, Major 32.5%, Mortality 15.8% | 328.5 days (iQR 83.8 - 666.8) |
|  |  | Stent (gastric outlet & colon) | 57 | 87.7% | 78.9% - Symptom relief | Median 4.5 days (iQR 2-11) | - | Reobstruction 38.6% | 30d: All grades 10.5%, Major 5.3%, Mortality 15.4% | 113 days (iQR 45.8 - 363.0) |
| Chen et al, 2020 (China) | GI (CRC & gastric) | Ostomy/bypass surgery (OBS) | 31 | - | 61.3% - Oral solid intake | - | 12.9% | Reobstruction 6.5% | 30d: All grades 29.0%, Major 0.0%, Mortality 19.4% | 5.4 months |
|  |  | Massive debulking surgery (MDS) | 30 | - | 90.0% - Oral solid intake | - | 20.0% | Reobstruction 3.3% | 30d: All grades 50.0%, Major 3.3%, Mortality 6.7% | 10.9 months |
| SINGLE ARM |  |  |  |  |  |  |  |  |  |  |
| Lodoli et al, 2021 (Italy) | Multiple (GI, Gyn, & others) | Surgery (stoma/resection/bypass/d ebulking/adhesiolysis) | 98 | 77.6% | 86.8% - Oral intake* | Median 16.5 days (iQR 10.4-25.8) | - | - | All grades 21.4%, Major 6.1%, In-hospital mortality 3.1% | - |
| De Boer et al, 2019 (Netherlands) | Multiple (GI, Gyn, & others) | Surgery for acute MBO (stoma/resection/bypass) | 148 | - | 83.7% - Oral intake** | Median 10 days (iQR 6-14) | 39.8%** | Reobstruction 35.0%, Readmission 56.1%** | All grades 58.1%, Major 20.9%, In-hospital mortality 8.8% | 119 days (iQR 48-420) |
| Zhang et al, 2022 (USA) | Multiple (GI, Gyn, & others) | Palliative CRS-HIPEC for MBO/ascites | 30 MBO (52 net) | - | 100% - Symptom relief | Median 11 days (iQR 9-13)^ | 69.2%^ | Reobstruction 10.7% | All grades 17.3%, Major 9.6%, Perioperative mortality 0.0%^ | 14.2 months (95% CI 3.7 - 20.5)^ |
| Baumgartner et al, 2019 (USA) | Appendix cancer | Surgery (stoma/resection/bypass/d ebulking/adhesiolysis +/- gastrostomy) | 26 | 100% | 96.2% - Return of Bowel Function + Oral intake | Median 14 days (range 5-36) | 34.6% | Readmission 23.1% for reobstruction | 60d major 23.1%, 30d mortality 3.8% | 18.5 months (95% CI 3.6-33.3) |
| Blair et al, 2001 (USA) | GI & Pancreatico-biliary | Surgery (stoma/resection/bypass/g astrostomy) | 63 | - | 46.0% - Oral solid intake | Median 12 days (range 3-39) | - | - | All grades 44.4%, including 30d mortality 20.6% | 3 months |
| Wong JSM et al, 2021 (Singapore) | Multiple (GI, Gyn, & others) | Surgery (GI/visceral resection/adhesiolysis +/- anastomosis +/- stoma) | 254 | - | - | - | - | Readmission 20.1% | 30d: All grades 43.3%, Major 24.8% including mortality 20.8% | 109 days (range 43-265) |
| Kawabata et al, 2022 (Japan - Multicenter) | Gastric cancer | Surgery (Resection/bypass +/- stoma) | 60 | - | 90.0% - Oral intake | Median 24 days (range 8-83) | 66.7% | - | All grades 44.4%, Major 15.9%, 30d mortality 3.2% | 6.6 months (95% CI 4.8–10.3) |
| Spiliotis et al, 2022 (Greece) | Multiple (GI, Gyn, & others) | Surgery (Resection/Anastomosis +/- debulking) | 300 | - | - | - | - | - | 30d: All grades 21.0%, Mortality 2.7% | 26.4 months |
| Higashi et al, 2003 (Japan) | Colorectal | Surgery (Resection/bypass/stoma) | 21 | - | - | - | 33.3% | - | 0% | Approx. 100 days |

Abbreviations: GI – Gastrointestinal, Gyn – Gynecologic, MBO – Malignant Bowel Obstruction, iQR – Interquartile Range, CD – Clavien Dindo Grade, CI – Confidence Interval
\*Oral intake was possible in 86.8% (66/76) of patients with surgical palliation compared to 4.5% (1/22) of patients who underwent exploratory laparotomy alone
\*\* Clinical success was defined for 104 patients who were discharged home and had data regarding dietary intake available; Chemotherapy initiation, Reobstruction, and Readmission data were reported for 123 patients who were discharged home
^Length of stay, chemotherapy initiation, complications and survival were reported for all included patients (with obstruction and ascites)

**Table 4:**
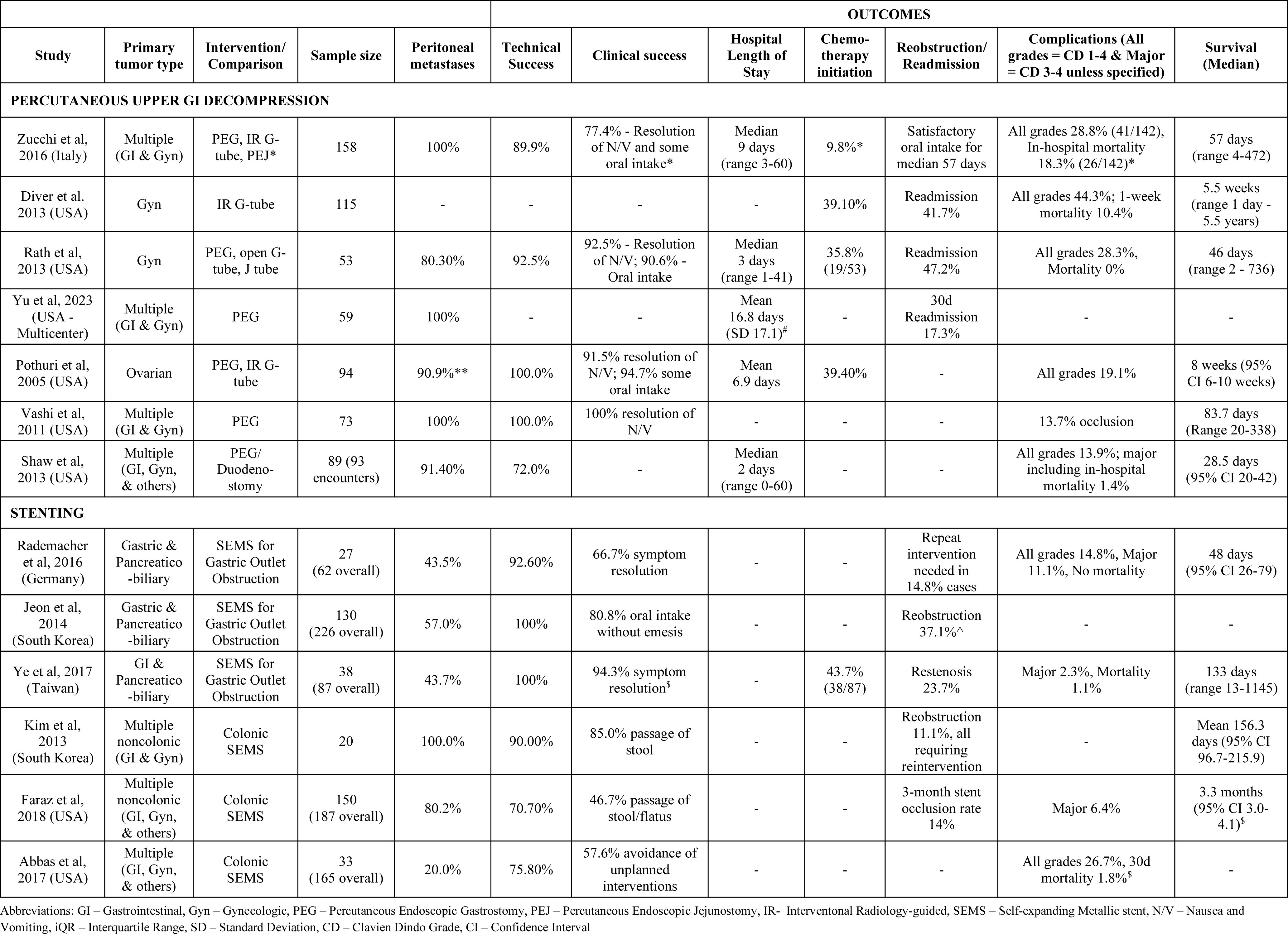
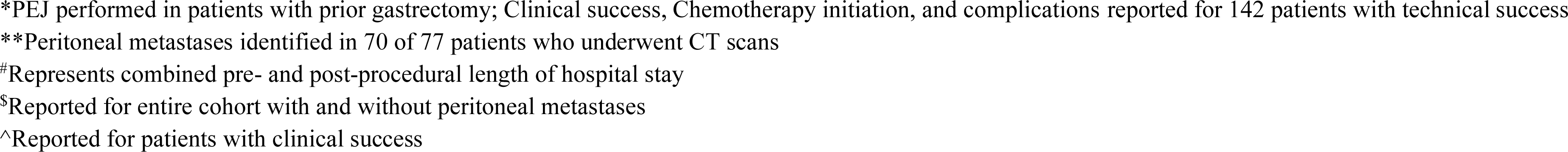
Summary of included studies focusing on percutaneous upper GI decompression tube placement and stenting.

|  |  |  |  |  | OUTCOMES |  |  |  |  |  |  |
| --- | --- | --- | --- | --- | --- | --- | --- | --- | --- | --- | --- |
| Study | Primary tumor type | Intervention/ Comparison | Sample size | Peritoneal metastases | Technical Success | Clinical success | Hospital Length of Stay | Chemo-therapy initiation | Reobstruction/ Readmission | Complications (All grades = CD 1-4 & Major = CD 3-4 unless specified) | Survival (Median) |
| **PERCUTANEOUS UPPER GI DECOMPRESSION** |  |  |  |  |  |  |  |  |  |  |  |
| Zucchi et al, 2016 (Italy) | Multiple (GI & Gyn) | PEG, IR G-tube, PEJ* | 158 | 100% | 89.9% | 77.4% - Resolution of N/V and some oral intake* | Median 9 days (range 3-60) | 9.8%* | Satisfactory oral intake for median 57 days | All grades 28.8% (41/142), In-hospital mortality 18.3% (26/142)* | 57 days (range 4-472) |
| Diver et al. 2013 (USA) | Gyn | IR G-tube | 115 | - | - | - | - | 39.10% | Readmission 41.7% | All grades 44.3%; 1-week mortality 10.4% | 5.5 weeks (range 1 day - 5.5 years) |
| Rath et al, 2013 (USA) | Gyn | PEG, open G-tube, J tube | 53 | 80.30% | 92.5% | 92.5% - Resolution of N/V; 90.6% - Oral intake | Median 3 days (range 1-41) | 35.8% (19/53) | Readmission 47.2% | All grades 28.3%, Mortality 0% | 46 days (range 2 - 736) |
| Yu et al, 2023 (USA - Multicenter) | Multiple (GI & Gyn) | PEG | 59 | 100% | - | - | Mean 16.8 days (SD 17.1)<sup>#</sup> | - | 30d Readmission 17.3% | - | - |
| Pothuri et al, 2005 (USA) | Ovarian | PEG, IR G-tube | 94 | 90.9%** | 100.0% | 91.5% resolution of N/V; 94.7% some oral intake | Mean 6.9 days | 39.40% | - | All grades 19.1% | 8 weeks (95% CI 6-10 weeks) |
| Vashi et al, 2011 (USA) | Multiple (GI & Gyn) | PEG | 73 | 100% | 100.0% | 100% resolution of N/V | - | - | - | 13.7% occlusion | 83.7 days (Range 20-338) |
| Shaw et al, 2013 (USA) | Multiple (GI, Gyn, & others) | PEG/ Duodeno-stomy | 89 (93 encounters) | 91.40% | 72.0% | - | Median 2 days (range 0-60) | - | - | All grades 13.9%; major including in-hospital mortality 1.4% | 28.5 days (95% CI 20-42) |
| **STENTING** |  |  |  |  |  |  |  |  |  |  |  |
| Rademacher et al, 2016 (Germany) | Gastric & Pancreatico -biliary | SEMS for Gastric Outlet Obstruction | 27 (62 overall) | 43.5% | 92.60% | 66.7% symptom resolution | - | - | Repeat intervention needed in 14.8% cases | All grades 14.8%, Major 11.1%, No mortality | 48 days (95% CI 26-79) |
| Jeon et al, 2014 (South Korea) | Gastric & Pancreatico -biliary | SEMS for Gastric Outlet Obstruction | 130 (226 overall) | 57.0% | 100% | 80.8% oral intake without emesis | - | - | Reobstruction 37.1%<sup>^</sup> | - | - |
| Ye et al, 2017 (Taiwan) | GI & Pancreatico -biliary | SEMS for Gastric Outlet Obstruction | 38 (87 overall) | 43.7% | 100% | 94.3% symptom resolution<sup>s</sup> | - | 43.7% (38/87) | Restenosis 23.7% | Major 2.3%, Mortality 1.1% | 133 days (range 13-1145) |
| Kim et al, 2013 (South Korea) | Multiple noncolonic (GI & Gyn) | Colonic SEMS | 20 | 100.0% | 90.00% | 85.0% passage of stool | - | - | Reobstruction 11.1%, all requiring reintervention | - | Mean 156.3 days (95% CI 96.7-215.9) |
| Faraz et al, 2018 (USA) | Multiple noncolonic (GI, Gyn, & others) | Colonic SEMS | 150 (187 overall) | 80.2% | 70.70% | 46.7% passage of stool/flatus | - | - | 3-month stent occlusion rate 14% | Major 6.4% | 3.3 months (95% CI 3.0-4.1)<sup>s</sup> |
| Abbas et al, 2017 (USA) | Multiple (GI, Gyn, & others) | Colonic SEMS | 33 (165 overall) | 20.0% | 75.80% | 57.6% avoidance of unplanned interventions | - | - | - | All grades 26.7%, 30d mortality 1.8%<sup>s</sup> | - |

Abbreviations: GI – Gastrointestinal, Gyn – Gynecologic, PEG – Percutaneous Endoscopic Gastrostomy, PEJ – Percutaneous Endoscopic Jejunostomy, IR- Interventonal Radiology-guided, SEMS – Self-expanding Metallic stent, N/V – Nausea and Vomiting, iQR – Interquartile Range, SD – Standard Deviation, CD – Clavien Dindo Grade, CI – Confidence Interval
\*PEJ performed in patients with prior gastrectomy; Clinical success, Chemotherapy initiation, and complications reported for 142 patients with technical success
\*\*Peritoneal metastases identified in 70 of 77 patients who underwent CT scans
#Represents combined pre- and post-procedural length of hospital stay
§Reported for entire cohort with and without peritoneal metastases

**Table 5:**
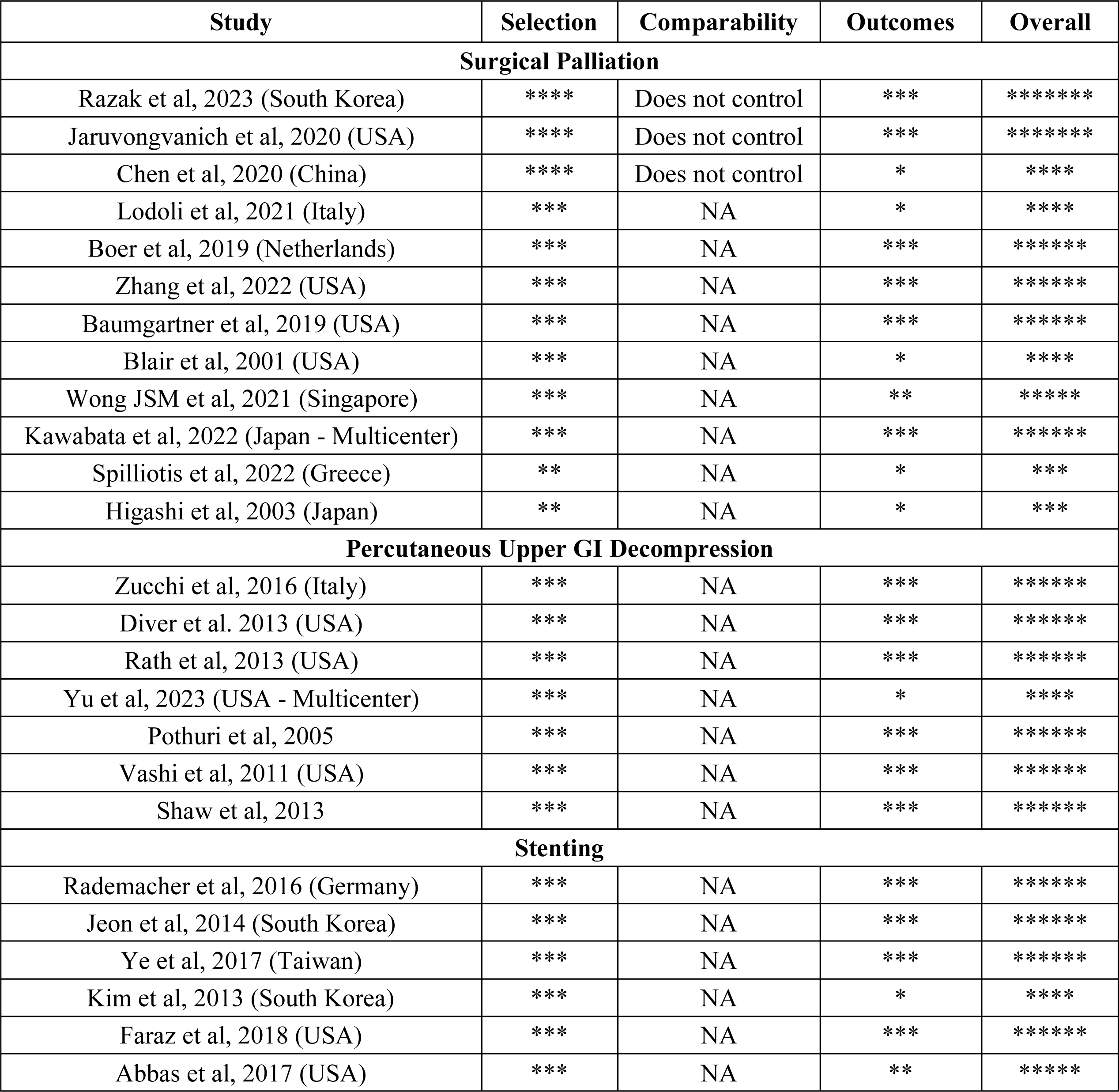
Quality assessments for included studies using the Newcastle Ottawa Scale for Cohort Studies.

#### Block 1 (Agreement: Round I - 97% Round II – 98%)

Block 1 outlines a comprehensive multidisciplinary approach for the initial workup, aligning with the general standard of care for bowel obstruction evaluation. Even in the setting of time constraints for high-acuity cases, prioritizing patient goals of care is essential. The workup includes a thorough oncologic history and physical exam, evaluation of cancer burden and prognosis, and assessment of prior surgical and non-surgical treatments. Standard laboratory panels and cross-sectional imaging of the abdomen and pelvis are integral components.

Cross-sectional imaging should be utilized to identify obstruction sites, particularly distinguishing small bowel obstructions, which are typically not eligible to manage with stents, from gastroduodenal and large bowel obstruction.^34^ An assessment of the focality of obstruction is imperative, with multifocal obstructions presenting greater technical complexity and poor prognosis with procedural interventions.^3,14,19,51^ Various studies in our review identified ascites as a negative prognostic factor for oral intake and survival after operative and non-operative interventions.^3,4,14,17,40,52^ Technical challenges in percutaneous endoscopic gastrostomy (PEG) tube placement can be anticipated in the presence of ascites and diffuse peritoneal metastases involving the greater omentum and gastric serosa.^53,54^

Resuscitation with crystalloids and bowel rest (nil per os status) is recommended, and nasogastric tube placement should be performed to decompress significant gastric distension and/or symptomatic nausea and vomiting, unless contraindicated. The initial patient interview should include discussions on overall goals of care, present quality of life, performance status, nutritional status, prognosis, and treatment options available with multidisciplinary input.^41^ Discussions about advance directives are crucial and should focus on the patient’s desires regarding their surrogate decision maker, cardiopulmonary resuscitation, mechanical ventilation, artificial nutrition, and hydration. Acknowledging the potential for fluctuations in goals of care throughout the course of treatment, the expert panel placed emphasis on iterative discussions, as highlighted in the top-most box in the pathway.^8,55^ Early specialist palliative care consultation is encouraged to facilitate multidisciplinary management of symptoms and supportive care needs across the patient’s disease course.^56–58^ For patients with subacute presentations such as chronic or recurrent bowel obstruction, consideration of discussion at a multidisciplinary tumor board is proposed when possible.

#### Block 2 (Agreement: Round I – 81%, Round II – 96%)

For patients presenting with an acute abdomen, i.e. findings on examination consistent with bowel perforation, necrosis, or peritonitis, a thorough discussion between the clinician, patient, and surrogate decision makers is necessary to clarify goals of care. While emergency surgery may be appropriate for those prioritizing prolongation of life, patients with a high disease burden or surgical risk may derive better quality of life from best supportive care and allowing natural death. Our review identified a study by de Boer et al. involving 148 patients with peritoneal carcinomatosis undergoing surgery for acute obstruction.^37^ Despite 83.1% of patients being discharged home postoperatively, 30% experienced severe postoperative complications (Clavien Dindo Grade 3-5) and 35% of discharged patients experienced recurrent obstruction.^37^ These results emphasize the substantial morbidity and mortality risks associated with emergency surgery for MGIO, underscoring the importance of goals of care discussions. In the first delphi round, this block did not emphasize the importance of these discussions adequately, yielding low consensus initially, which improved in round II after added emphasis.

#### Block 3 (Agreement: Round I - 94%, Round II – 98%)

The choice between upfront primary medical management and surgical intervention for patients without an urgent surgical indication has been a matter of equipoise. Historical evidence from observational series within and beyond the scope of our review (before January 2000) favored definitive surgical intervention when feasible, demonstrating benefits in restoring oral intake, improving obstructive symptoms, facilitating systemic chemotherapy initiation, and prolonging life. ^2,13,59–61,33^ However, these series likely exhibit selection bias favoring surgical cohorts with lower age, better performance status, and a lower burden of peritoneal carcinomatosis. Our review identified SWOG S1316 as the first prospective trial comparing surgery vs. non-surgical management for malignant small bowel obstruction.^12^ This trial included 199 patients, of which 66.3% had suspicion or evidence of peritoneal carcinomatosis. The primary outcome was the mean number of “good days”, i.e. days alive and spent out of the hospital, in the first 13 weeks after enrollment, which showed no significant difference between surgery and non-surgical management (adjusted mean difference 2.9 days, p = 0.50). Overall survival at 13 weeks (HR 0.70 (95% CI 045-1.09), p = 0.12) and oral intake at 5 weeks (> 80 % in both groups) also did not differ significantly between the two treatment groups. Prespecified exploratory analyses revealed no significant associations between the number of good days and overall survival with the presence of peritoneal carcinomatosis, although subgroup analyses were not reported. Despite some treatment-related complications in patients treated with surgery (15% vs. 4% in the non-surgical management group), they had improved GI symptom scores. These results suggest that surgically eligible patients may derive benefit in health-related quality of life with an operation early in their hospital stay, although it does not significantly impact good days, overall survival, or oral intake.

For patients without an indication for urgent surgery, attempting a trial of medical management is recommended. The 2018 Chicago Consensus Guidelines highlight various approaches, including combinations of nasogastric decompression, intravenous hydration, and medications for controlling symptoms and gastric secretions such as somatostatin analogues, corticosteroids, histamine-2 antagonists, and proton pump inhibitors.^27^ Individualization based on multidisciplinary discussions is crucial due to the lack of strong evidence supporting a specific medical therapy for improving MGIO resolution rate, time, or survival. Since there is insufficient evidence specific to patients with peritoneal carcinomatosis, recommendations are extrapolated from the broader MGIO evidence spectrum. A 2016 systematic review encompassing seven randomized controlled trials on somatostatin analogues included two placebo-controlled trials with low-risk of bias, both of which found no significant symptomatic improvement with somatostatin analogues in their primary end points.^62–64^ However, one of these, a trial with 80 patients with peritoneal carcinomatosis, demonstrated a reduction in vomiting episodes and improved patient well-being with Lanreotide vs. placebo in secondary analyses.^64^ Corticosteroids, evaluated in a Cochrane review including three randomized controlled trials, exhibited a trend suggesting potential resolution of bowel obstruction with intravenous dexamethasone, although statistical significance was not reached.^65^ Oral water soluble contrasts such as gastrografin lack sufficient evidence of benefit in MGIO, with a 2018 Cochrane review identifying only a pilot RCT involving nine patients comparing gastrografin to placebo.^66,67^ Hence, their potential use is left to the discretion of the treating physician.

If the patient’s clinical status fails to improve or worsens with up to one week of medical management, it may constitute persistent obstruction, necessitating surgical intervention if feasible and within the patient’s goals of care.^33^ Upfront PGID placement can also be discussed, especially if anatomical considerations or recurrent presentations preclude durable symptom improvement with medical management alone.

#### Block 4 (Agreement: Round I – 97%, Round II – 97%)

When faced with persistent MGIO, reassessing the patient’s clinical status and goals is imperative when determining their eligibility for surgery. Factors known to be associated with diminished surgical benefit include high peritoneal disease burden, multifocal obstruction, ascites, multiple metastatic sites, high-grade histology, poor performance status, and indicators of poor nutrition, such as low serum albumin. ^3,4,12,14,33,35,40,41,44,52,68,69^ Small bowel obstruction is associated with a poor prognosis, as highlighted in Lodoli et al.’s study including 98 patients with peritoneal carcinomatosis. Amongst these patients, definitive surgery was not possible in 22% of cases, indicative of technical failure.^36^ Colonic obstructions, followed by ileal, and jejunal or more proximal obstructions were associated with an incremental likelihood of failure; mesenteric involvement and retraction were also associated with failure. Additionally, three other studies revealed that small bowel obstruction was associated with lower rates of oral intake and survival postoperatively compared to obstructions not involving the small bowel.^14,24,40^ The decision to proceed with surgery should carefully consider these factors, weighing benefits against morbidity in the context of limited remaining life expectancy, which may range from 1-6 months in the presence of high-risk features.^5,12,14,33,70^ Alternative options for patients deemed unsuitable for definitive surgery include PGID and best supportive care with or without hospice referral.

### Percutaneous Upper GI Decompression (PGID)

Decompressive gastrostomy tubes, primarily placed endoscopically but also through interventional radiology (IR) or surgery, are placed with the intention of alleviating upper GI symptoms such as nausea, vomiting, and bloating, and enable limited, palliative oral intake for the patient’s remaining life. In individuals with prior gastrectomy, decompressive jejunostomy or duodenostomy may be considered. Our review identified seven studies on PGID in patients ineligible for definitive surgery (Tables 4 and 5).^7–9,45–48^

The technical success rate ranged from 72.0% to 100%,^8,9,45,47,48^ with clinical success variably defined as the resolution of symptoms or oral intake, reaching rates from 77.4% to 100%.^8,9,45,47^ Median post-procedure hospital stay duration spanned 2 to 9 days, ^8,9,45,48^ and chemotherapy initiation post-decompression, observed in 9.8% to 39.4% of patients, correlated with improved survival in select reports (Diver et al, 2013 and Rath et al, 2013).^7–9,45^ Complication rates varied from 13.7% to 44.3%, including peristomal infection (3.2% - 14.1%), leakage (1.4% - 8.5%), tube dislodgement (2.7% - 13.9%), and obstruction (3.2% - 17.0%), necessitating repositioning or replacement in 0.7% to 17.0% of cases.^7–9,45,47,48^ Post-procedure readmission rates ranged from 17.3% to 47.2%,^7,8,46^ and median post-procedure survival durations ranged from 28.5 days to 83.7 days, emphasizing symptomatic relief rather than prolonging survival as the primary purpose of PGID placement.^7–9,13,45,47,48^

In the presence of ascites, PGID placement can pose challenges but remains feasible. Rath et al. observed that in 53 patients with gynecological malignancies, 38 had ascites, with three requiring paracenteses before percutaneous gastrostomy tube placement.^8^ Although PEG could not be completed initially in four patients with ascites due to inadequate transillumination, successful placement was achieved on the second attempt through various methods (PEG under general anesthesia/open surgical placement/IR-jejunostomy). In another study by Pothuri et al., all 94 patients with recurrent ovarian cancer achieved technical success of PEG, with 25 requiring paracenteses before the procedure.^9^ Shaw et al. detailed 93 gastrostomy encounters in patients with malignant bowel obstruction and ascites, the latter being managed simultaneously by paracentesis (13 cases) or intraperitoneal catheters (78 cases).^48^ Compared to other studies with relatively fewer patients with ascites, they reported lower technical success rates (72/3, 77.4%) and higher rates of major complications (7/72, 9.7%), including peritonitis in four patients. Percutaneous transesophageal gastrostomy (PTEG) serves as an alternative in cases with ascites and diffuse gastric involvement.^53,54,71,72^

#### Block 5 (Agreement: Round I – 86%, Round II – 94%)

For patients considering surgery or stenting as treatment options for MGIO, the approach is determined by the site of obstruction and desired outcomes, whether immediate relief or a pathway to definitive therapy. In a study by Jaruvongvanich et al which compared outcomes of stenting (upper GI or colon) and surgical correction (stoma/resection/bypass) for MGIO due to peritoneal carcinomatosis, baseline differences such as better performance status favored the surgery group.^34^ The surgery group also presented with more small bowel and multifocal obstructions, whereas the stent group had more colon and gastric outlet obstructions, which were more accessible endoscopically. While the surgery group exhibited higher rates of symptom resolution (95.0% vs. 78.9%, p = 0.03) and a trend toward improved survival (median 328.5 days vs. 113 days, p = 0.06), they also experienced longer hospital stays (median 9 days vs. 4.5 days, p = 0.008) and high 30-day major morbidity rates (32.5% vs. 5.3%, p < 0.001). While this retrospective series did not adjust for baseline differences between the two cohorts, it underscores the importance of obstruction location and patient performance status in treatment selection. Extensive discussion of the risks and benefits of each approach should occur on an individualized basis.

### Surgery

In our review encompassing 12 studies on surgical palliation for MGIO in patients with peritoneal carcinomatosis, eight studies addressed surgical correction, three focused on tumor debulking alongside surgical correction, and one addressed acute abdominal emergencies as detailed in Block 2 (Tables 3 and 5).^33–44^ Technical success was defined as the ability to perform surgical correction, with rates ranging from 77.6% to 100%.^33,36,39^ One study reported 100% technical success, however six patients with non-therapeutic laparotomy were excluded, suggesting an anticipated technical success rate below 100%.^39^ Clinical success, denoting symptomatic relief or return of bowel function, ranged from 61.3% to 100%, except for a study by Blair et al, reporting a low clinical success rate of 46.0%.^33–36,38–40,42^ Hospital stays averaged between 9 and 24 days,^33,34,36–40,42^ while postoperative chemotherapy was initiated in 12.9% to 69.2% of cases.^33,35,37–39,42,44^ Re-obstruction rates varied from 3.3% to 47.5%,^33–35,38,39,41^ with postoperative complication rates between 17.3% and 50.0%.^33–36,38–43^ Major complications, graded Clavien Dindo 3-4 equivalent, occurred in 0% to 24.8% of cases and were attributed mainly to intra-abdominal or cardiopulmonary sequelae such as collections requiring drainage, sepsis, bleeding, myocardial infarction, pulmonary embolism, and respiratory distress.^33–36,38,39,41,42^ Average overall survival was 3-11 months for surgical correction studies,^33–36,39–42^ and ranged from 11 to 26 months for debulking studies, suggesting a highly selected cohort in the latter. ^35,38,43^

The least invasive surgical approach should be pursued for obstructive relief. Bypass or stoma creation may be considered if resection/anastomosis is precluded by the extent or location of obstructions, mesenteric foreshortening, or concern for short gut. Comparative evidence between different surgical correction methods is limited, relying on retrospective studies involving diverse patient populations beyond peritoneal carcinomatosis. For instance, Perri et al. found differences in overall survival in 62 patients with recurrent gynecologic malignancies undergoing surgery for MBO: resection-anastomosis/bypass vs. colostomy alone vs. ileostomy alone (median 270 days vs. 154 days vs. 82 days, p < 0.03).^68^ Similarly, Legendre et al. highlighted superior clinical success and survival with resection over bypass.^16^ PGID placement may be included with any surgical approach for reliable enteral access or as an option for symptom relief if other avenues are exhausted.

### Stenting

In cases of endoscopically accessible obstructions like gastric outlet or colonic obstructions, self-expanding metallic stents (SEMS) placement may be viable. Our review included six studies regarding stenting in patients with peritoneal carcinomatosis which reported technical and clinical success rates ranging from 70.7% to 100% and 46.7% to 94.3%, respectively (Tables 4 and 5).^17,18,23,49–51^ Factors associated with clinical success included unifocal obstructions compared to multifocal obstruction and the absence of ascites.^17,51^ Obstructions with lengths exceeding 4 cm may necessitate the placement of multiple stents and carry a higher chance of failure, as highlighted in a study including patients with and without peritoneal carcinomatosis.^20^ Re-obstruction or stenosis rate ranged from 11.1% to 37.1% and was reported to occur at a median duration of 8 to 16 weeks from initial stent placement, often requiring repeat intervention.^17,18,49–51^ Major complications (Clavien-Dindo 3-4 equivalent) were reported in 2.3% to 11.1% of cases, with bowel perforation and gastrointestinal bleeding being the most common stent-related complications, whereas stent migration was infrequently reported. ^49,51,73^ During the consensus, a recommendation was made to consider diverting ostomy instead of stenting for rectal obstructions owing to high risks of post-procedure pain and perforation with the latter. Stenting in jejuno-ileal obstructions is less common but has shown feasibility in limited series via percutaneous jejunostomy under IR guidance.^74^ Although not mentioned explicitly in the pathway, experts in the consensus suggested the consideration for endoscopic lumen apposing metallic stent (LAMS)-assisted gastro-jejunal or entero-enteric bypass for gastric outlet or more distal obstructions, respectively, as an alternative option for palliation.^75–77^

The block version in round I lacked clarity regarding the selection of definitive surgical interventions and SEMS and yielded low consensus initially. These were elucidated in the second round with subsequent improvement in consensus levels. When stenting is not feasible, surgical exploration should be considered via the least invasive safe approach, often beginning with diagnostic laparoscopy. Biopsies should be obtained intraoperatively if additional tissue is needed to assess the underlying malignancy, even if no further resections are planned. If intraoperative findings indicate that further intervention is unsafe or unfeasible, surgery should be concluded, and management decisions should align with the strategies outlined in Block 4.

#### Block 6 (Agreement: Round I – 93%, Round II – 98%)

If resolution of the bowel obstruction is obtained, the focus shifts to preventing recurrence of obstruction and enhancing quality of life. Continuous assessment of performance status, nutritional health, pathology, and care goals, including advance directives, is crucial. A palliative care consult should be considered with a focus on symptom management for all patients throughout their disease course. It may also be combined with curative-intent treatment, in contrast with modes of care at terminal stages of life, such as hospice. Systemic chemotherapy or clinical trials may be considered in select patients who can tolerate further treatment. Best supportive care utilizing hospice, with consideration of palliative nutrition and hydration, may help provide nourishment for the remainder of life. The appropriateness of parenteral nutrition in MGIO remains uncertain, as highlighted in a 2018 Cochrane Review.^78^ Current recommendations align with the 2018 Chicago Consensus Guideline, suggesting consideration of parenteral nutrition in patients eligible for standard chemotherapy, possessing satisfactory performance status (ECOG =< 2 / KPS > 50%), and predicted survival of at least two months.^27^ Social support and psychological preparedness should be evaluated before initiating parenteral nutrition, as it is a demanding course of treatment. Cytoreduction with or without intraperitoneal chemotherapy may be considered for surgical candidates, whereas hospice evaluation is recommended for individuals unlikely to benefit from aggressive interventions.

## DISCUSSION

The current effort represents a substantial advancement over the Chicago Consensus Guidelines 2018 as it establishes a formal pathway with supporting evidence for procedural interventions tailored to patients with peritoneal carcinomatosis.^27^ Over 95% consensus was achieved across all six blocks after two rounds of review by a multidisciplinary group. This methodology aims to provide clarity regarding clinical management despite issues highlighted in prior reviews surrounding the heterogeneity and quality of available evidence.^1,28,79,80^ Our guideline also includes recommendations for gastric and duodenal obstruction unlike prior reviews that primarily focused on MBO (i.e., obstruction distal to the ligament of Treitz).^28,80^ Furthermore, our literature search reflects advancements in medical and surgical management over the last two decades.

It is essential to recognize several limitations inherent to our review and consensus when interpreting the recommendations. Our review primarily focused on procedural interventions, although evidence concerning medical management was derived from included studies comparing surgical and medical approaches where applicable, supplemented by extrapolation from existing systematic reviews.^62,65,66,78^ The studies encompassed various primary tumor sites (gastrointestinal, gynecological, and others), each with unique biology and potential impacts on outcomes. Moreover, there was variability in the proportion of patients with peritoneal carcinomatosis among the included studies. To mitigate this, summary tables were confined to studies reporting any peritoneal carcinomatosis, despite variations in the definition of peritoneal carcinomatosis across studies and uncertainty regarding its role as the cause of obstruction in all cases.

Additionally, success rates with procedural interventions may be overrepresented in highly selected cohorts excluding patients with high-grade or high-burden peritoneal disease upfront. The variability in definitions and time points for outcome measurements further limited our ability to pool outcome statistics, compounded by the lack of adequate comparative evidence controlling for baseline differences. Quality of life-related outcomes were infrequently measured and highly subjective, with only two out of 62 studies reporting the use of validated quality of life instruments during follow-up.^12,42^ Furthermore, the limitations of the consensus process should be acknowledged, particularly the majority composition of the expert panel consisting of surgical oncologists. Having expected this bias from during the inception phases, experts in specialist palliative care, medical oncology, and nutrition were involved early on for reviewing feedback from the first Delphi round. Voting on blocks rather than individual itemized recommendations was preferred to align with the original Chicago Consensus framework and to prevent survey fatigue, although this approach may compromise the granularity of feedback received and over represent consensus percentages.

Despite these limitations, the guideline delineates best practices from experts regarding the management of MGIO in patients with peritoneal carcinomatosis, offering a framework to standardize clinical care and reduce variation. The consensus underscores several key areas of equipoise regarding the sequence and selection criteria for medical and procedural interventions, along with strong agreement on the importance of evaluating and aligning with goals of care throughout treatment. While these results provide valuable guidance for clinical practice and treatment decisions, they do not replace the need for high-quality evidence concerning this vulnerable patient population.

### External perspectives

Our methodology and recommendations align with existing guidelines for managing MGIO in other expert consensus documents, as well as societies outside of the United States. A joint guideline from French societies across disciplines focusing on peritoneal carcinomatosis presents a decision tree outlining indications and selection criteria for surgery and stenting, similar to our pathway.^79^ Additionally, they propose a three-stage medical management protocol in the absence of surgical options and emphasize the importance of venting gastrostomy versus long-term nasogastric tube placement for persistent intractable vomiting. Other guidelines, while not specifically tailored to patients with peritoneal carcinomatosis, provide valuable insights. The Multinational Association for Supportive Care in Cancer offers comprehensive recommendations for MBO care, with a substantial focus on medical management and nutrition.^81^ In contrast, our panel involves more surgical oncologists, highlighting potential differences in emphasis and expertise.

The NCCN Guidelines for Palliative Care offer a closely aligned algorithm for assessing and managing malignant bowel obstruction, stressing the importance of treatment decisions based on goals of care discussions, patient education, and psychosocial support.^82^ Similarly, surgical guidelines from Northern America and Europe addressing the role of endoscopy in managing MGIO identify analogous criteria for SEMS insertion.^83–85^ Notably, the recommendation for surgical gastrojejunostomy over self-expanding metallic stents (SEMS) for patients with malignant gastric outlet obstruction (GOO) and a life expectancy over two months differs from our findings. This recommendation is based on studies primarily involving patients with GOO of locoregional origin,^86–88^ which may not be applicable to our focus on patients with peritoneal carcinomatosis, highlighting the need for context-specific guidelines tailored to distinct patient populations.

## Conclusion

This document describes a clinical pathway for managing MIGO in patients with PSM, developed through a modified Delphi consensus including surgical oncologists, medical oncologists, and palliative care specialists. The pathway encourages specialist palliative care assessments and continuous evaluation of goals of care throughout treatment. Recommendations advocate aligning treatment decisions regarding medical or procedural interventions with individual goals of care in emergent and non-emergent scenarios. A rapid review of evidence revealed limited benefits of palliative surgery and stenting in patients with multifocal obstructions, poor performance status, and high-grade or high-burden PSMs. In such situations, supportive care or the placement of upper GI decompression tubes was preferred.

## Supporting information

Supplementary figure - Pathway round 1 version

Rapid review search strategy

## Data Availability

The datasets used and/or analysed during the current study are available from the corresponding author on reasonable request.

