## Supplementary figure - Pathway round 1 version for "Consensus Guideline for the Management of Malignant Gastrointestinal Obstruction in Patients with Peritoneal Surface Malignancies"

### Malignant Bowel Obstruction due to peritoneal carcinomatosis

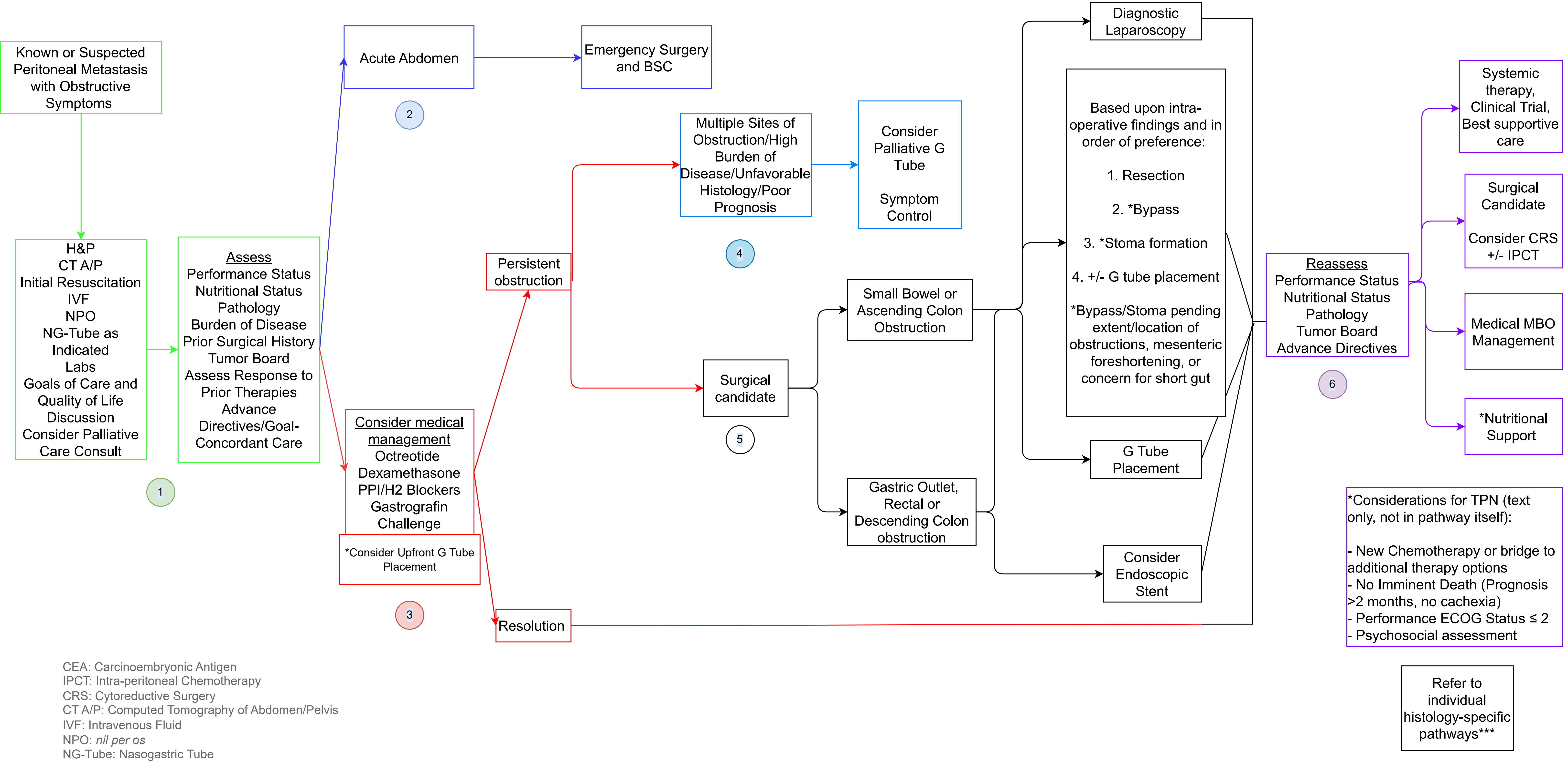


CEA: Carcinoembryonic Antigen
IPCT: Intra-peritoneal Chemotherapy
CRS: Cytoreductive Surgery
CT A/P: Computed Tomography of Abdomen/Pelvis
IVF: Intravenous Fluid
NPO: *nil per os*
NG-Tube: Nasogastric Tube
BSC: Best Supportive Care
PPI: Proton Pump Inhibitor
MBO: Malignant Bowel Obstruction
