## Supplementary material for "Consensus Guideline for the Management of Malignant Gastrointestinal Obstruction in Patients with Peritoneal Surface Malignancies": Rapid review search strategy

Search Strategy for rapid review.

| | |
|---|---|
| **22** | #20 AND #21 |
| **21** | periton*[tw] OR carcinomatosis*[tw] OR *unresect[tw] OR incurable*[tw] OR carcinosis*[tw] OR un-resect*[tw] |
| **20** | #11 AND #19 |
| **19** | #14 AND #18 |
| **18** | #15 OR #16 OR #17 |
| **17** | Resect*[tw] OR Surger*[tw] OR surgical*[tw] OR Debulk*[tw] OR Bypass*[tw] OR ileostom*[tw] OR colostom*[tw] OR gastrostom*[tw] OR stoma*[tw] OR ("Surgical stomas"[Mesh Major Topic]) OR cytoreduct*[tw] OR ("Cytoreduction surgical procedures"[Mesh Major Topic]) |
| **16** | Surgical Procedures, Elective[MeSH Major Topic] |
| **15** | Colorectal surgery[MeSH Major Topic] |
| **14** | #12 OR #13 |
| **13** | (bowel*[tw] or intestin*[tw] or gastrointestin*[tw] or gastro-intestin*[tw] or colon*[tw] or colorect*[tw] or retrosigmoid*[tw]) AND (obstruct*[tw] or blockage*[tw]) |
| **12** | intestinal obstruction[MeSH Major Topic] |
| **11** | #1 OR #2 OR #3 OR #10 |
| **10** | #6 AND #9 |
| **9** | #7 OR #8 |
| **8** | neoplasm*[tw] OR cancer*[tw] OR carcinoma*[tw] OR tumor*[tw] OR tumour*[tw] |
| **7** | Neoplasms[MeSH Major Topic] |
| **6** | #4 OR #5 |
| **5** | gastrointestinal*[tw] OR gastro-intestin*[tw] OR intestin*[tw] OR bowel*[tw] OR colon*[tw] OR colorectal*[tw] OR rectal*[tw] OR stomach*[tw] or gastric*[tw] |
| **4** | gynecologic*[tw] OR gynecologic*[tw] OR ovar*[tw] OR endometr*[tw] OR uter*[tw] OR vagina*[tw] OR vulvar*[tw] OR cervi*[tw] |
| **3** | Ovarian Neoplasms |
| **2** | Genital Neoplasms, Female[MeSH Major Topic] |
| **1** | Gastrointestinal neoplasms[MeSH Major Topic] |
